# An Open-Label Feasibility Study of Remdesivir in Long COVID: Results from the ERASE LC Trial

**DOI:** 10.64898/2026.07.26.26358852

**Authors:** Mark A Faghy, Jade Chynoweth, Anton Barnett, Lindsay Skipper, Victoria Allgar, Helen Neilens, Kayle-Anne Sands, Amber Lord, Helen Hambly, Paigan J Aspinall, Chris Rollinson, W David Strain, Rebecca Owen, Callum Thomas, Marie J Grigg, Sascha H Kranen, Kinan Mokbel, Karen M Knapp, Hairil Abdul Razak, Ruth Ashton, Ian Maidment, Tom Bewick

**Affiliations:** School of Sport, Exercise and Health Sciences, Loughborough University, Loughborough, UK; College of Health and Humanities, University of Derby, Derby, UK; Medical Statistics Research Group, University of Plymouth, Plymouth, UK; Peninsula Clinical Trials Unit, University of Plymouth, Plymouth, UK; Faculty of Health and Life Sciences, University of Exeter, Exeter, UK; Research Centre for Healthcare and Communities Transformation, Coventry University, UK; College of Health and Life Sciences, Aston University, Birmingham, UK; University Hospitals of Derby and Burton NHS Foundation Trust, Derby, UK

**Keywords:** Long COVID, feasibility study, viral persistence, anti-viral treatment, public health

## Abstract

**Objective:** Treatment of acute COVID-19 with anti-viral and immunomodulator medications demonstrates a reduced risk of long-term outcomes. To date, remdesivir, an intravenous (IV) antiviral that prevents RNA transcription, has not been evaluated in people with Long COVID (LC). This study assessed the feasibility of a five-day IV remdesivir intervention for individuals with LC.

**Methods:** Seventy-three participants aged ≥18 years with a confirmed LC diagnosis were recruited across two sites in the United Kingdom to receive a five-day course of IV remdesivir. Primary feasibility outcomes included recruitment, treatment completion, acceptability, and safety. Exploratory patient-reported (e.g. Fatigue Assessment Scale [FAS]) and clinical outcomes (e.g. 6-minute walk test [6MWT]) were also collected.

**Results:** Of 106 individuals screened, 73 were enrolled and 71 (97%) completed the 5-day IV remdesivir regimen. Follow-up assessments were completed by 70 participants (96%), with high completion rates observed across clinical assessments, patient-reported outcomes and symptom tracking. No severe adverse reactions were reported. Improvements were also observed in several exploratory patient-reported and clinical outcomes.

**Conclusion:** We demonstrated the feasibility and acceptability of administering IV remdesivir in people with LC. Analysis of secondary outcomes suggests therapeutic promise, but rigorous evaluation in large-scale randomised controlled trials is essential to determine the true efficacy and clinical utility of intravenous remdesivir in this population.

**Lay Summary:** Long COVID (LC) can cause ongoing symptoms like fatigue and reduced physical ability after a COVID-19 infection. Some medicines used during acute COVID-19 infections, such as antivirals, may lower the risk of long-term problems, but it isn’t clear whether they can help people who already have Long COVID.

This study looked at whether it is practical and safe to give remdesivir (an antiviral drug given through a drip in the arm) to people with Long COVID over five days.

A total of 73 adults with Long COVID took part across two sites in the UK. Almost all participants (97%) were able to complete the full treatment. About one-third experienced side effects, but none were serious. Researchers also saw signs of improvement in symptoms like fatigue and in physical performance tests, although these were not the main focus of the study.

Overall, the study shows that giving remdesivir to people with Long COVID is feasible and generally well tolerated. While early results suggest it might help, larger and more rigorous studies are needed to find out if it truly works.

## Introduction

Post-acute sequelae of SARS-CoV-2 infection, also termed post-COVID-19 condition or Long COVID (LC), is a heterogeneous multisystem disorder characterised by symptoms lasting at least three months after acute infection, without an alternative explanation [1]. Global prevalence remains difficult to ascertain due to inconsistent diagnostic criteria and reduced surveillance, but current estimates suggest that LC affects between 65 and 400 million people worldwide [2,3]. Distinct subtypes of LC have been described. Some result from direct viral injury to cardiovascular or neurological systems, while others reflect sequelae proportional to acute illness severity, such as post-ICU syndromes. One of the most debilitating presentations is the myalgic encephalomyelitis/chronic fatigue syndrome (ME/CFS) phenotype, characterised by disproportionate fatigue, unrefreshing sleep, cognitive dysfunction (also known as ‘brain fog’), and post-exertional malaise (PEM, [4] PEM complicates the assessment and treatment, as even minimal physical, cognitive, orthostatic and emotional activity can trigger exacerbated symptoms. In research settings, PEM can be quantified using two-day cardiopulmonary exercise testing (CPET), where individuals show reduced function on the second test despite identical exertion [5]. Although advances have been made in characterising the clinical spectrum, epidemiology, and pathophysiology of LC, its underlying mechanisms remain incompletely defined [6]. Current evidence supports multiple interacting processes, including persistent viral reservoirs, dysregulated immunity, autoimmunity, latent virus reactivation, endothelial and microvascular dysfunction, microclots, gut dysbiosis, and impaired mitochondrial metabolism [7,8]. Neurobiological studies additionally implicate chronic neuroinflammation, microglial activation, disrupted neurovascular integrity, and altered synaptic remodelling, contributing to cognitive impairment, sensory symptoms, and fatigue [9].

Viral persistence, whether through ongoing low-level replication, residual viral genomes, or delayed antigen clearance, has been hypothesised as a contributing factor in the pathology of LC [8]. Persistent SARS-CoV-2 antigens and ribonucleic acid (RNA) have been detected across tissues even after resolution of acute symptoms, supporting the premise that a subset of individuals with LC are not able to fully clear viral material [10,11]. Similar persistent viral remnants are well recognised in other single-stranded RNA infections, including Ebola, Zika, enteroviruses, and measles [7,12]. Persistent SARS-CoV-2 proteins, microthrombi, and inflammatory signatures have also been reported in tissues and post-mortem samples, reinforcing the biological plausibility of sustained viral reservoirs [13–18].

The role of antiviral therapeutics in LC is driven by accumulating evidence that persistent viral reservoirs or viral products may contribute to ongoing symptomatology.[8] Antiviral therapy, whether specific for SARS-CoV-2 with nirmatrelvir (administered with ritonavir to improve bioavailability, marketed as Paxlovid) or repurposed from other disease areas (e.g., molnupiravir, tenofovir disoproxil/emtricitabine (TDF/FTC) or maraviroc), when administered in acute SARS-CoV-2 have reduced the incidence of LC [19–22]. To date, however, oral therapies have failed to demonstrate clinically meaningful benefits in LC, even after extended dosing strategies [19,23].

Remdesivir is an IV nucleotide analogue prodrug that acts by inhibiting the viral RNA-dependent RNA polymerase [24]. This posology allows for higher intracellular concentrations over a longer duration than can be achieved with oral formulations. It does, however, require consecutive days of treatment, which exerts additional limitations on a population living with chronic and debilitating symptoms, including post-exertional malaise. We therefore sought to determine the recruitment, retention and tolerability of a trial using this treatment regimen. Trial feasibility and the determination of progression to a definitive trial were the primary outcomes. To appropriately power the definitive clinical trial, we collected clinically relevant patient-reported outcome measures and clinical assessments for descriptive analysis.

## Methods

### Study Design

ERASE-LC was a two-centre (Derby and Exeter, UK), open-label, single-arm feasibility trial evaluating the deliverability, acceptability, and tolerability of a five-day IV remdesivir regimen in adults with LC. The full protocol, schedule of assessments, and prespecified progression (‘stop–go’) criteria have been published previously [25]. The trial was registered on relevant trial registries (ClinicalTrials.gov NCT05911906; ISRCTN 72940450). The study was conducted in accordance with Good Clinical Practice and reported in line with Consolidated Standards of Reporting Trials (CONSORT) guidance for pilot and feasibility trials. Ethics approval was obtained from the UK Health Research Authority Research Ethics Committee (24/SC/0118), the Administration of Radioactive Substances Advisory Committee (AA-8765), and the Medicines and Healthcare products Regulatory Agency (24/SC/0118). All participants provided written informed consent before any study procedures.

### Participants

Eligible participants were adults (≥18 years) with clinician-confirmed LC (post-COVID-19 condition) and previous confirmed or suspected SARS-CoV-2 infection, with persistent symptoms relative to pre-COVID baseline as captured by patient-reported outcomes. Additional inclusion criteria were the ability to provide informed consent, complete study questionnaires and planned clinical assessments, attend scheduled visits, and live within commutable distance of the study site (at the discretion of the local principal investigator). Full criteria are provided in the protocol [25]. Key exclusion criteria were prior remdesivir, nirmatrelvir/ritonavir, molnupiravir, or other COVID-19 antiviral use within the preceding 6 months; known immunocompromise; current participation in a structured physical rehabilitation programme aimed at LC; inability to complete procedures due to severe risk of PEM as judged clinically by the Modified DePaul Symptom Questionnaire (DSQ-PEM, 26], pregnancy, breastfeeding, or attempting to conceive; unable to understand verbal English/have a hearing impairment that prevents adequate communication, participation in another clinical drug trial within the last 6 months, taking concomitant medications with known clinically significant interactions with remdesivir; history of serious infusion reactions; and clinically significant hepatic or renal impairment.

### Patient and Public Involvement and Engagement (PPIE)

People with lived experience of LC contributed to all aspects of the trial, and this was achieved through an established PPIE group. Involvement was critical at all stages of the study, including review of participant-facing materials and developing study and site-specific safety mitigations (including reducing or eliminating the risk of PEM, and exposure to airborne and fomite pathogens) whilst undertaking all aspects of the trial. PPIE representatives were core members of the trial leadership team and had representation in trial oversight structures (e.g., Trial Management Group and Trial Steering Committee), ensuring that the study maintained a patient-centred focus.

### Intervention and safety monitoring

Participants attended their local site on five consecutive days for IV remdesivir. Visits included assessment of adverse events since prior contact, infusion administration, and post-infusion observation. Participants able to conceive underwent pregnancy testing at each dosing visit. Remdesivir was administered per the Summary of Product Characteristics: a 200 mg loading dose on day 1 (IV infusion over 60 minutes), followed by 100 mg on days 2–5 (IV infusion over 30 minutes), each in 250 mL 0·9% sodium chloride. Participants were observed for at least 30 minutes after each infusion. Predefined stopping criteria included clinically significant infusion reactions or other safety concerns judged by the treating clinician; participants discontinuing treatment were withdrawn from further dosing but were invited to complete follow-up assessments where feasible.

### Procedures and assessments

Assessments were completed at baseline and post-intervention follow-up (approximately 28 days after completion of dosing). Patient-reported outcome measures (PROMs) were completed electronically (REDCap) with paper alternatives available as a reasonable adjustment. PROMs included fatigue (Fatigue Assessment Scale [FAS] and Modified Fatigue Impact Scale [MFIS]), health-related quality of life (EQ-5D-5L), symptom burden (Symptom Burden Questionnaire for LC [SBQ™-LC]), functional status (Post-COVID Functional Status scale), dyspnoea (Medical Research Council dyspnoea scale), cognitive symptoms (Perceived Deficit Questionnaire [PDQ-5]), generalized anxiety disorder questionnaire (GAD-7) and PEM (DSQ-PEM). Clinical assessments included the six-minute walk test (6MWT) with Borg perceived exertion and oxygen saturation, venous blood sampling for inflammatory and metabolic biomarkers, and a two-day CPET protocol (Exeter site only) to quantify exercise capacity and exertional responses (full details in protocol [25]. Participants also undertook daily symptom tracking using a smartphone application (Visible Health, UK), where possible and those without smartphone access used paper diaries. Additional symptom-tracking and wearable-monitoring and physiologic data were collected as part of the wider feasibility programme; detailed analyses of these datasets are beyond the scope of the present manuscript and will be reported separately.

### Outcomes

The primary outcomes were feasibility and acceptability metrics: recruitment rate, consent rate, retention and follow-up completion, adherence to the five-day dosing regimen, and completeness of key study assessments (PROMs, clinical measures, biomarkers, and CPET). Safety outcomes included adverse events and adverse reactions. Further secondary/exploratory outcomes included changes from baseline in PROMs, functional capacity measures (6MWT), day-to-day CPET tolerability, variables, and selected biomarkers, intended to inform outcome selection and sample size planning for a definitive randomised controlled trial.

### Statistical Analysis

Participant-reported and clinical outcomes were summarised according to the intention-to-treat principle (i.e. including all participants regardless of adherence to the study drug). This feasibility trial was not powered to support or justify any conclusions regarding treatment efficacy/effectiveness deduced from any hypothesis testing. The analysis did not include formal inferential statistical comparisons or hypothesis testing. All analyses were undertaken once the final participant had completed their final assessment, and the database was locked. Baseline characteristics collected during screening after consent were summarised descriptively by recruitment site to provide an overview of the trial population. Continuous data were summarised by mean and standard deviation (SD), unless data were at least moderately skewed, in which case, the median and interquartile range were used. Categorical variables were summarised by frequencies and percentages of non-missing responses. Pre-post intervention differences were summarised and presented with two-sided 75%, 85% and 95% confidence intervals (CIs). Analyses were adjusted for site.

#### Role of the Funding Source

The funder has had no role in the design, delivery, interpretation and dissemination of the data/results of this study. Funding was provided on a researcher-led funding agreement.

## Results

### Recruitment and Retention

A total of 106 participants were identified between September 2024 and May 2025. Thirty-three participants were excluded (20 ineligible, 8 too busy, 3 general health reasons, 1 uncontactable and 1 personal circumstances). Of the 20 participants deemed ineligible, 11 (55%) were excluded because they were identified as being at high risk of severe post-exertional malaise (PEM) following engagement in physical tasks, as determined using the DePaul Symptom Questionnaire. The remaining exclusions were due to logistical, clinical, or eligibility-related reasons, including inability to attend study visits, participation in concurrent rehabilitation programmes, compromised immune function, history of serious adverse reactions to medications or infusions, inability to provide informed consent, or absence of a confirmed Long COVID diagnosis. Seventy-three participants provided consent and were recruited. There were two withdrawals during the study, one withdrew consent before baseline assessments and one after day 2 of the IV remdesivir treatment due to an adverse reaction. One further participant was lost to follow-up at post-intervention assessment due to other unrelated health reasons. A further five participants did not complete all post-intervention CPET assessments. A CONSORT diagram is shown in Figure 1.

**Figure 1:**
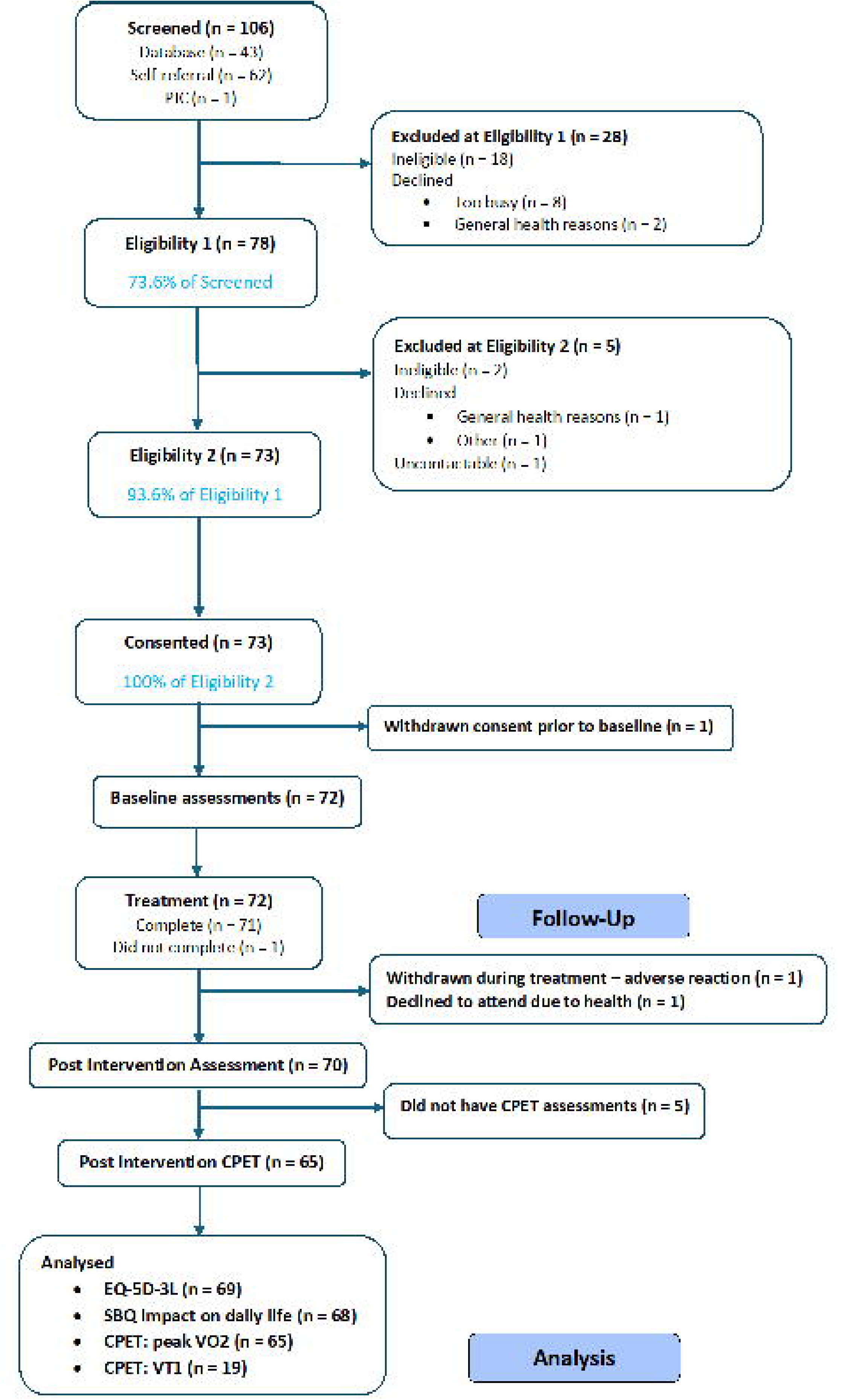
CONSORT-style diagram of participant flow through the ERASE-LC feasibility study A diagram depicting the flow of participants through each stage of the recruitment process.

### Demographics & Descriptive Characteristics

The baseline characteristics and demographics are shown in Table 1.

**Table 1:** Participant baseline characteristics and demographics.

|  |  | Derby (n=53) | Exeter (n=20) | All (n=73) |
| --- | --- | --- | --- | --- |
| **Age (years)** |  |  |  |  |
| n: mean (SD) [min, max] |  | 52: 50.1 (11.3) [26.0, 72.0] | 20: 49.0 (13.2) [19.0, 80.0] | 72: 49.8 (11.8) [19.0, 80.0] |
| median (Q1, Q3) |  | 50.5 (43.0, 58.0) | 47.0 (43.5, 56.5) | 49.5 (43.0, 57.5) |
| Missing |  | 1 (1.9) | 0 (0.0) | 1 (1.4) |
| **Sex** | Male | 22 (42.3) | 5 (25.0) | 27 (37.5) |
|  | Female | 30 (57.7) | 15 (75.0) | 45 (62.5) |
|  | Missing | 1 (1.9) | 0 (0.0) | 1 (1.4) |
| **Ethnicity** | White | 49 (94.2) | 20 (100.0) | 69 (95.8) |
|  | Mixed/Multiple | 0 (0.0) | 0 (0.0) | 0 (0.0) |
|  | Asian<sup>1</sup> | 2 (3.8) | 0 (0.0) | 2 (2.8) |
|  | Black<sup>2</sup> | 1 (1.9) | 0 (0.0) | 1 (1.4) |
|  | Other | 0 (0.0) | 0 (0.0) | 0 (0.0) |
|  | Missing | 1 (1.9) | 0 (0.0) | 1 (1.4) |
| **BMI (kg/m<sup>2</sup>)** |  |  |  |  |
| n: mean (SD) [min, max] |  | 52: 29.2 (5.2) [21.3, 41.6] | 20: 26.2 (5.5) [19.4, 41.2] | 72: 28.4 (5.4) [19.4, 41.6] |
| median (Q1, Q3) |  | 28.5 (24.4, 34.1) | 25.1 (22.9, 28.2) | 27.1 (24.1, 31.7) |
| Missing |  | 1 (1.9) | 0 (0.0) | 1 (1.4) |
| **Rate health prior to COVID-19** | Very good | 35 (67.3) | 16 (80.0) | 51 (70.8) |
|  | Good | 15 (28.8) | 3 (15.0) | 18 (25.0) |
|  | Moderate | 2 (3.8) | 1 (5.0) | 3 (4.2) |
|  | Bad | 0 (0.0) | 0 (0.0) | 0 (0.0) |
|  | Very bad | 0 (0.0) | 0 (0.0) | 0 (0.0) |
|  | Missing | 1 (1.9) | 0 (0.0) | 1 (1.4) |
| **Rate health today** | Very good | 0 (0.0) | 0 (0.0) | 0 (0.0) |
|  | Good | 2 (3.8) | 1 (5.0) | 3 (4.2) |
|  | Moderate | 12 (23.1) | 5 (25.0) | 17 (23.6) |
|  | Bad | 28 (53.8) | 12 (60.0) | 40 (55.6) |
|  | Very bad | 10 (19.2) | 2 (10.0) | 12 (16.7) |
|  | Missing | 1 (1.9) | 0 (0.0) | 1 (1.4) |
| **Number of confirmed COVID infections** |  |  |  |  |
| n: mean (SD) [min, max] |  | 52: 2.1 (1.0) [0.0, 4.0] | 20: 2.4 (1.1) [1.0, 5.0] | 72: 2.2 (1.0) [0.0, 5.0] |
| median (Q1, Q3) |  | 2.0 (1.0, 3.0) | 2.0 (1.5, 3.0) | 2.0 (1.0, 3.0) |
| Missing |  | 1 (1.9) | 0 (0.0) | 1 (1.4) |
| **Number of suspected COVID infections** |  |  |  |  |
| n: mean (SD) [min, max] |  | 52: 2.5 (1.2) [1.0, 6.0] | 20: 2.7 (1.4) [1.0, 5.0] | 72: 2.6 (1.2) [1.0, 6.0] |
| median (Q1, Q3) |  | 2.0 (2.0, 3.0) | 2.5 (1.5, 4.0) | 2.0 (2.0, 3.0) |
| Missing |  | 1 (1.9) | 0 (0.0) | 1 (1.4) |
| **Number of**<br>**COVID vaccines**<br>**received** | 0 | 0 (0.0) | 0 (0.0) | 0 (0.0) |
|  | 1 | 0 (0.0) | 1 (5.0) | 1 (1.4) |
|  | 2 | 6 (11.5) | 1 (5.0) | 7 (9.7) |
|  | 3 | 12 (23.1) | 6 (30.0) | 18 (25.0) |
|  | 4 | 13 (25.0) | 3 (15.0) | 16 (22.2) |
|  | 5+ | 21 (40.4) | 9 (45.0) | 30 (41.7) |
|  | Missing | 1 (1.9) | 0 (0.0) | 1 (1.4) |

The mean (SD) age of participants was 49.8 (11.8) years. There was a larger proportion of females (75%) at the Exeter site. Overall, 69 (96%) participants in the study were of white ethnicity. The mean (SD) BMI of participants was 28.4 (5.4). Only 3 (4%) participants rated their health “today” as good or very good. Nearly all participants (67 – 93%) were referred to a LC clinic. The median (Q1, Q3) score for whether LC symptoms affected participants’ daily life was 8 (7, 10) – with 0 having no effect and 10 having a big effect, with the lowest score reported as 4. Overall, 53 (74%) participants had never smoked, and there were no current smokers; 33 (46%) participants had no comorbidities. Overall, 61 (84%) participants were on medication, and of those taking medication, the median (Q1, Q3) number of medications taken was 3 (2, 5).

### Feasibility Outcomes

A total of 106 participants were screened across two sites, and 73 consented to take part. Of the 43 participants self-referred through the database, 32 (74%) consented; an additional 62 self-referred via the website, with 40 (65%) consenting. In total, 73 participants enrolled, of whom 71 (97%) completed the full 5-day IV remdesivir regimen. Post-intervention assessments were completed by 70 participants (96%), and CPET follow-up was achieved in 65 participants (89%).

Completeness of clinical assessments exceeded 85% at both sites across all time points are highlighted in Table 2. Overall completeness at both baseline and post-intervention remained high for the biomarkers IL-1β, IL-4, IP-10 and RANTES (>80%) and moderate for IL-8 and IL-12 (>75%). Except for the SBQ™-LC, patient-reported outcome measures were completed by more than 90% of participants at each assessment. The SBQ™-LC was completed by 64% at baseline and 52% post-intervention, rising to 95% and 81%, respectively, when excluding the sexual health domain. Baseline CPET day 1 and 2 assessments were completed by 72 participants (99%), and post-intervention CPET assessments were completed by 65 participants (89%). Reasons for non-completion of post-intervention CPET assessments were post-exertional malaise (n=1), symptom exacerbation considered unrelated to study procedures (n=1), adverse events (n=1), travel burden (n=1), and unrelated personal factors (n=1), indicating that only one participant was unable to complete follow-up CPET due to PEM. At the PET/CT site, 17 participants (85%) completed baseline scans and 12 (60%) completed post-intervention scans. Overall, 69 participants (95%) completed at least one day of symptom tracking.

**Table 2:** Feasibility outcomes and progression against prespecified stop-go criteria.

| Feasibility outcome | Red | Amber | Green | Observed outcome | Progression status |
| --- | --- | --- | --- | --- | --- |
| Site delivery | No sites able to run study | 1 of 2 sites able to run study | 2 of 2 sites able to run study | Both sites successfully delivered the study and completed recruitment | Green |
| Recruitment uptake (proportion recruited once eligible) | <25% | 25–50% | >50% | 73/73 eligible participants recruited (100%) | Green |
| Treatment fidelity (completion of all 5 remdesivir infusions) | <70% | 60–85% | >85% | 71/73 participants completed treatment (97%) | Green |
| Follow-up completion (28 days post-IMP) | <65% | 65–85% | >85% | 70/73 participants completed follow-up assessments (96%) | Green |
| Completion of key outcome measures | <60% | 60–80% | >80% | Clinical assessment completion >85%; most PROMs >90%; key biomarker completion >80% | Green |
| Completion of CPET assessments | <60% | 60–80% | >80% | 65/73 participants completed post-intervention CPET assessments (89%) | Green |
| Safety and tolerability* | N/A | N/A | Acceptable safety profile | 47 adverse reactions reported by 22 participants (31%); none were severe | Acceptable |

### Safety Data

There was one serious adverse event during the study, assessed as unrelated to the intervention by the local investigator. A total of 64 adverse events, including 47 adverse reactions, occurred in 26 participants (36%), with 22 participants (31%) experiencing adverse reactions. Of the 47 adverse reactions, none were severe, and 43 (91%) were mild (16 headaches, 9 other, 3 nausea, 3 influenza-like illness, 3 fatigue, 3 migraine with aura, 2 migraine (without aura), 2 flank pain, 2 abnormal liver test). The highest number reported by any single participant was five (four mild and one moderate). The timing of adverse events was also examined in relation to study procedures. Of the 47 adverse reactions classified as related to the intervention, 43 (91%) occurred during the five-day remdesivir infusion period, with the remaining four occurring after the safety blood check. No treatment-related adverse reactions were reported following CPET assessments, PET/CT imaging, or post-intervention outcome assessments, suggesting that the majority of adverse reactions were temporally associated with treatment administration rather than the exercise-based assessments.

### Key Patient Reported, Biomarker and CPET Outcomes

Table 3 presents the pre/post intervention differences and confidence intervals for the predefined key outcomes, adjusted for recruitment site.

**Table 3.** Adjusted pre/post intervention differences and confidence intervals in key outcome measures.

| Measure |  | n at baseline | n at follow up | Pre/post intervention change | 75% confidence interval | 85% confidence interval | 95% confidence interval |
| --- | --- | --- | --- | --- | --- | --- | --- |
| EQ-5D utility index score (mapped to EQ-5D-3L) |  | 72 | 69 | 0.060 | [0.036, 0.085] | [0.029, 0.091] | [0.018, 0.102] |
| SBQ™-LC – impact on daily life subtotal |  | 72 | 68 | -11.228 | [-13.661, -8.795] | [-14.273, -8.183] | [-15.374, -7.082] |
| Biomarkers | IL-1β | 68 | 64 | -0.249 | [-2.095, 1.596] | [-2.559, 2.060] | [-3.394, 2.895] |
|  | IL-4 | 67 | 63 | 40.766 | [2.057, 79.474] | [-7.674, 89.205] | [-25.187, 106.718] |
|  | IL-8 | 57 | 55 | 12.670 | [1.194, 24.145] | [-1.691, 27.030] | [-6.883, 32.222] |
|  | IL-12 | 61 | 57 | 2.980 | [0.295, 5.664] | [-0.380, 6.339] | [-1.595, 7.554] |
|  | IP-10 | 65 | 60 | 1.671 | [0.395, 2.948] | [0.074, 3.269] | [-0.504, 3.846] |
|  | RANTES | 62 | 60 | -279.569 | [-585.457, 26.319] | [-662.353, 103.215] | [-800.741, 241.603] |
| CPET | VT1 | 28 | 33 | 8.991 | [4.862, 13.120] | [3.824, 14.157] | [1.956, 16.025] |
| | Peak $\dot{V}O_2$ | 72 | 65 | -0.025 | [-0.392, 0.341] | [-0.484, 0.434] | [-0.650, 0.600] |

Using a 95% confidence interval, the pre/post intervention differences in the patient-reported outcomes were: EQ-5D-5L index score (mapped to EQ-5D-3L;[27]: 0.06 (95% CI: 0.02, 0.10) and SBQ™-LC – impact on daily life subtotal: -11.23 (95% CI: -15.37, -7.08, Figure 2). For the CPET outcomes, using a 95% confidence interval, the pre/post intervention difference in the change (between day 1 and 2) in the first ventilatory threshold (VT1) was 8.99 work rate (watts) (95% CI: 1.96, 16.03). For the biomarker outcomes, using an 85% confidence interval, the pre/post intervention differences were: IP-10: 1.67 pg/mL (85% CI: 0.07, 3.27). Using a 75% confidence interval, the pre/post intervention differences were: IL-4 pg/mL: 40.77 (75% CI: 2.06, 79.47), IL-8: 12.67 pg/mL (75% CI: 1.19, 24.15) and IL-12: 2.98 pg/mL (75% CI: 0.30, 5.66).

**Figure 2:**
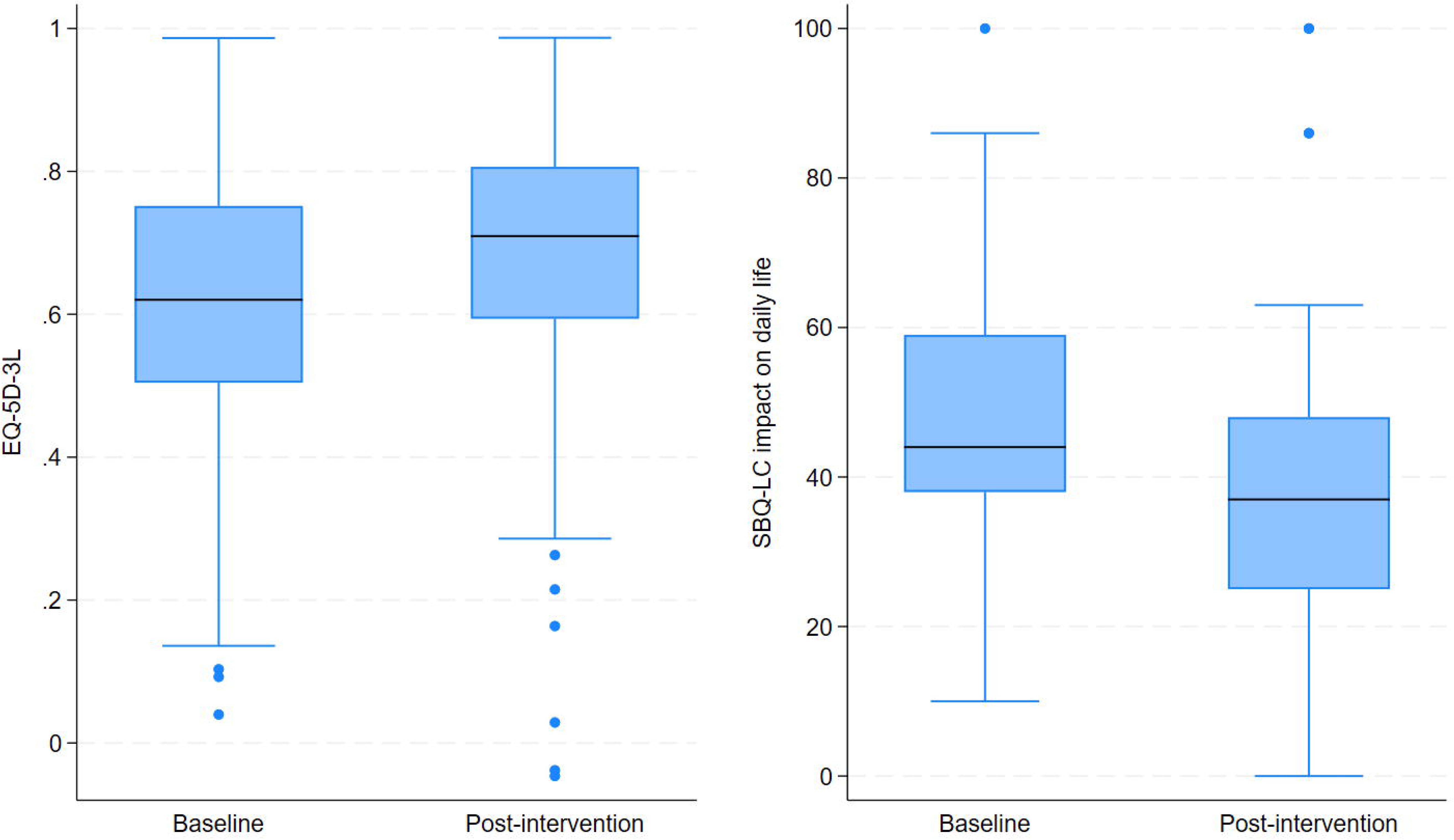
Box plots of EQ-5D-5L (mapped to EQ-5D-3L) utility score (left) and SBQ-LC impact on daily life (transformed score) (right) at baseline and post-intervention. A figure showing the changes in quality of life before and after the trial.

### Other Patient Reported and Physiological Outcomes

The mean (SD) of the FAS total score at baseline was 34.3 (7.3) compared to 28.3 (8.0) post-intervention (Figure 3). Overall, 53 (78%) participants had a reduction in the FAS total score, with 42 (62%) participants showing a reduction of at least 4 points. An additional 11 (15%) participants demonstrated improvement that did not reach the MCID threshold, whilst 3 (4%) participants remained unchanged and 12 (17%) demonstrated worsening fatigue scores. Among participants who did not improve, the mean (SD) increase in FAS score was 2.2 (1.4) points (range 0–4). Overall, 36 (50%) participants were rated as severe on the FAS at baseline compared to 14 (21%) post-intervention. Furthermore, 29 (42%) participants moved to a less fatigued FAS category, 38 (55%) remained in the same category, and only 2 (3%) moved to a more fatigued category. Both participants in the latter group moved from the normal to mild-to-moderate fatigue category, with the increase limited to 3 points in each case. The mean (SD) of the MFIS total score at baseline was 55.7 (12.9) compared with 42.4 (17.8) post-intervention (Figure 3). At baseline, 19 (26%) participants reported moderate or severe anxiety on the GAD-7 compared to 7 (10%) post-intervention. The mean (SD) distance achieved in the 6MWT was 408 (113) metres at baseline compared to 468 (109) metres post-intervention (Figure 4). The median (Q1, Q3) of the EQ-5D visual analogue score (VAS) at baseline was 50 (35, 61) compared to 60 (40, 71) post-intervention. At baseline, 53 (75%) participants had a moderate or severe post-COVID-19 functional status category compared to 33 (49%) post-intervention.

**Figure 3:**
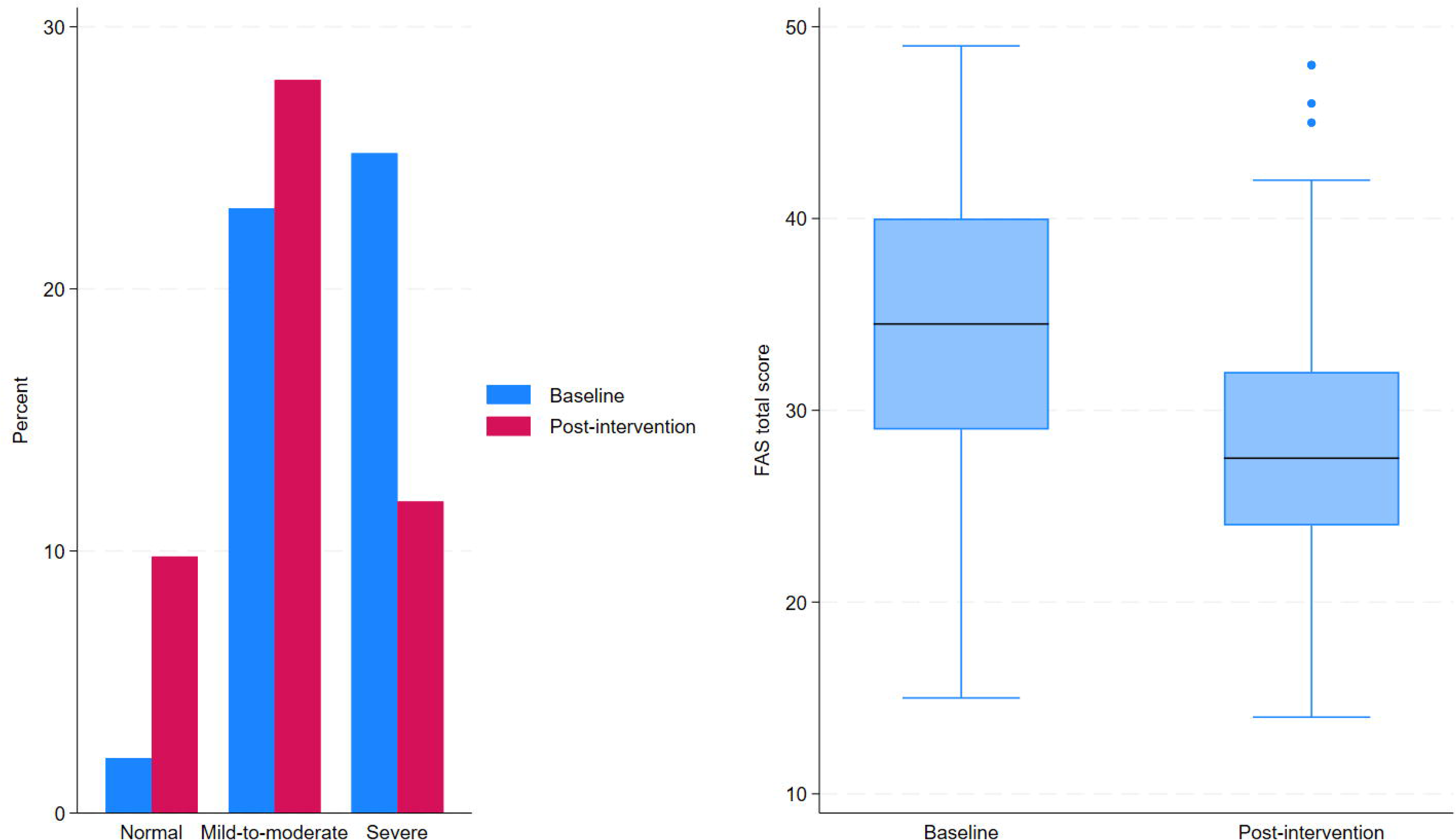
Box plot of FAS total score (left) and bar chart of FAS category (right) at baseline and post-intervention. A figure showing the changes in fatigue that patients reported before and after the trial.

**Figure 4:**
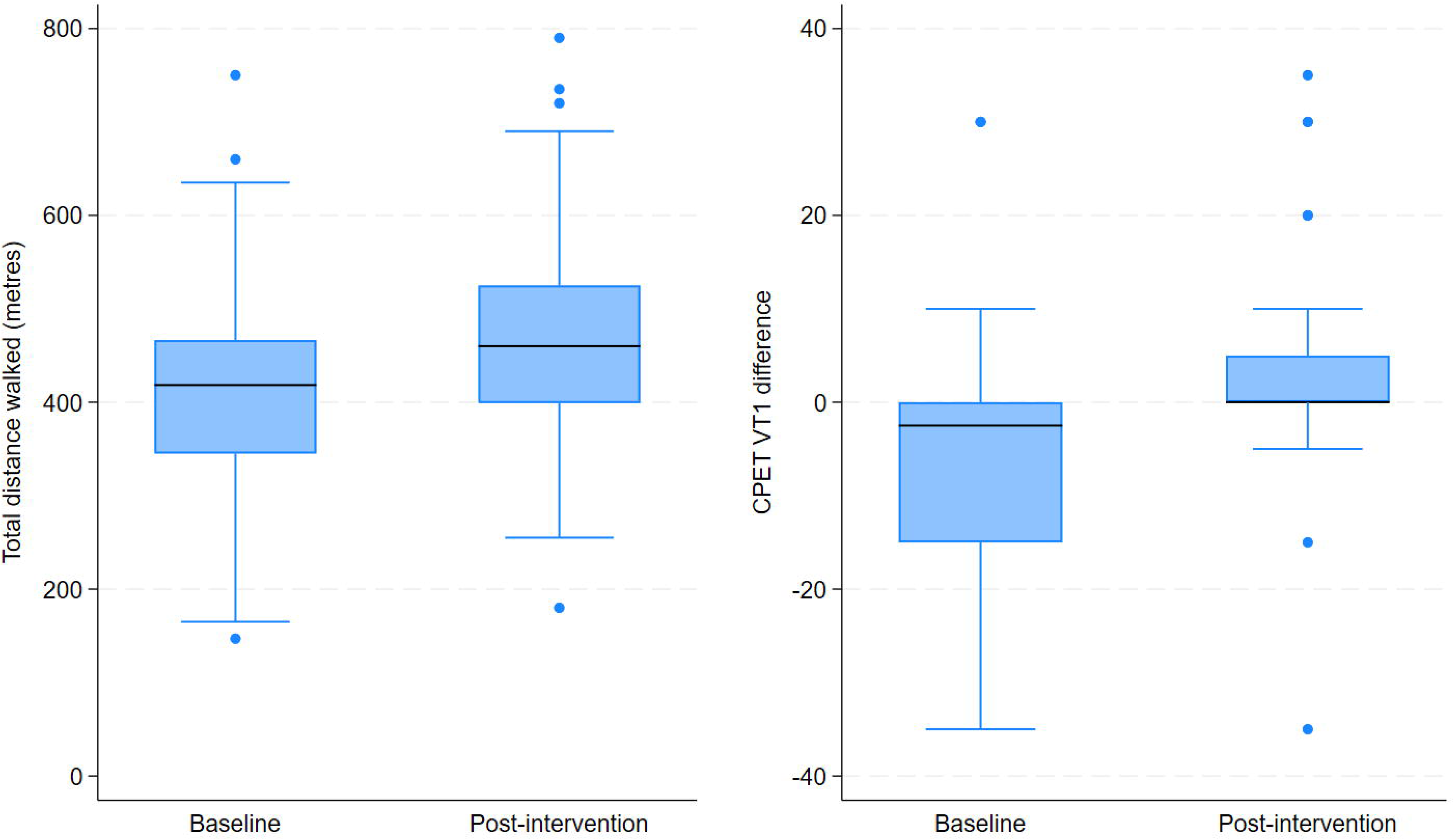
Box plots of 6-minute walk test (metres) (left) and CPET VT1 (right) showing between-day differences at baseline and post-intervention. A figure showing an improved distance covered on the study’s walking test that was completed by participants.

### Progression to a definitive randomised controlled trial

The progression to a definitive randomised controlled trial according to the ‘stop-go’ criteria can be found elsewhere [28] and in the protocol paper [25]. Both sites were able to run the study; those who were confirmed as eligible at the detailed screening visit was 100% from those marked eligible from screening, treatment fidelity (defined as completion of all five IV remdesivir sessions) was 97% and follow-up at 28 days post intervention was 96% (all rated as green). Completion of the key outcomes at baseline and post-intervention was >80% (rated as green) for each outcome except SBQ^TM –^LC, which was 64% and 52% at baseline and post-intervention, respectively (95% at baseline and 81% post-intervention if excluding the sexual health sub-scale) and the IL-8 and IL-12 biomarkers, which were 75% and 78% post-intervention, respectively.

## Discussion

The key findings from this trial indicate that recruiting to a trial administering remdesivir via IV infusion to people with mild-to-moderate LC is both feasible and safe in this cohort, with no observed or reported unexpected safety concerns. Analysis demonstrates an objective change in ventilatory threshold compatible with an improvement in symptoms, supporting the clinical suggestion to progress to the definitive trial. The descriptive analysis demonstrates numerical improvements of 6 points in the FAS total score (clinically meaningful being regarded as ≥3 points, [29] and various domains of quality-of-life scores, including symptom burden, quality of life, functional capacity, and physiological measures. Given that one objective of this feasibility study was to inform outcome selection for a future definitive trial, it is important to consider the clinical relevance of these observed changes. The improvement in FAS exceeded the established MCID [30], supporting its suitability as a candidate efficacy outcome in future studies. Similarly, the observed improvement in 6MWT distance exceeded published MCID estimates reported in other chronic disease populations (14.0–30.5 m) [31], suggesting a potentially meaningful improvement in functional capacity. In contrast, whilst improvements were observed in health-related quality of life, the mean change in EQ-5D-5L index score (0.06) did not exceed the published MCID of 0.11 [32], indicating that improvements in this measure should be interpreted more cautiously. Improvements were also observed in CPET measures, including the first ventilatory threshold, which provide objective physiological evidence that complements patient-reported outcomes. However, LC-specific MCIDs for CPET variables are not currently established, making interpretation of the clinical importance of these changes more challenging. CPET completion rates were high, with 89% of participants completing post-intervention assessments. Importantly, only one participant did not complete follow-up CPET because of post-exertional malaise. Other reasons for non-completion included symptom exacerbation considered unrelated to study procedures, an adverse event, travel burden, and unrelated personal factors. These findings provide reassurance regarding the tolerability of the CPET protocol in the selected study population. Nevertheless, the occurrence of PEM in one participant, alongside the exclusion of individuals considered at high risk of severe PEM, highlights the need to carefully balance the value of detailed physiological assessment against participant burden and inclusivity in future trials.

From an outcome-selection perspective, FAS appears particularly well suited for inclusion in a future definitive trial, demonstrating change beyond the established MCID and aligning with its increasing use as an outcome measure in LC [30,33]. The 6MWT also demonstrated potentially clinically meaningful change and provides an objective assessment of functional capacity. In contrast, EQ-5D-5L may be less sensitive to short-term treatment effects in this population given that the observed change did not exceed the published MCID. CPET outcomes provided valuable physiological insight and objective evidence of change; however, their role in future studies may be better suited to mechanistic evaluation or nested sub-studies rather than as mandatory assessments for all participants. Data completeness also informed outcome selection. Whilst completion rates exceeded 90% for most patient-reported outcome measures, completion of the sexual health domain within the SBQ™-LC was substantially lower. Completion rates improved markedly when this domain was excluded, suggesting that acceptability concerns may have contributed to missing data. Future studies should therefore consider whether inclusion of this domain is essential or whether a modified SBQ™-LC could improve acceptability and data completeness while retaining comprehensive assessment of symptom burden. It is important to note that these changes in patient-reported outcomes are unadjusted. Whilst these early findings are encouraging, they remain preliminary and confirmation via a rigorously designed and adequately powered randomised controlled trial is essential to determine the true efficacy, durability of effect, and cost-effectiveness of remdesivir before it can be recommended for widespread clinical use in the treatment and management of LC.

In the context of LC, this is the first trial to determine the feasibility of delivering antiviral medication over 5 days via IV infusion, is feasible and safe. At present, this must be compared with experience from other relevant clinical areas, such as acute infection with SARS-CoV-2 and community-acquired pneumonia, where IV therapies are standard. There are established protocols for the administration of monoclonal antibodies for infectious diseases, chemotherapy in oncology, biologic infusions in rheumatology, and parenteral antimicrobials in acute care, which provide valuable learning into the promise and the practical challenges associated with scaling IV-based treatments in both research and clinical practice [34]. Data from these areas illustrate that IV delivery can achieve high bioavailability, rapid therapeutic onset, and consistent dosing, all of which may be advantageous in addressing complex, multisystem conditions like LC [35]. Despite demonstrating feasibility, the scaling and implementation of IV treatment approaches will require careful planning, including adequate infusion-centre capacity, safety mitigations, which include reducing the risk of incidental infection/re-infection, exacerbating PEM, flexible appointments, accessibility and clear monitoring and safety protocols to address infusion-related reactions and ensure equitable access. Experience from established clinical pathways shows that although IV therapies can be successfully integrated into routine practice at scale, doing so depends on coordinated infrastructure, well-defined clinical guidelines, and thoughtful patient selection processes [36]. As larger trials and real-world evaluations emerge, these cross-disciplinary insights will be essential for determining how IV antiviral treatments can be delivered efficiently, safely, and equitably to broader LC populations.

Viral persistence is recognised as a significant pathology in LC [7,8]. Whilst our data is not conclusive, it provides encouraging early indications that treatment with remdesivir, delivered via IV methods, could be effective at improving patient outcomes for those living with LC. Parallel efforts within large research consortia, including the RECOVER clinical trials program [37] have embedded antiviral candidates within broader multimodal therapeutic studies designed to target fatigue, exertional intolerance, sleep disturbances, and neurocognitive impairment. To date, no antiviral has yet shown consistent, robust efficacy across diverse patient populations; indeed, several large randomised controlled trials of the MPro inhibitor nirmatrelvir, packaged with ritonavir as Paxlovid, have failed to show benefit [38]. Paxlovid, however, is dependent on actively replicating SARS-CoV-2. Although particles of viral RNA and SARS-CoV-2 antigen have been identified in people with LC, active viral replication has not been identified. Remdesivir is a direct genomic RNA synthesis inhibitor that impairs SARS-CoV-2 replication [39]. Administered IV, in a pro-drug form, it is then converted intracellularly into the active triphosphate, where it acts on viral RNA transcription. Importantly, however, it is not dependent on whole viral replication to be effective, but rather it inhibits RNA-dependent RNA polymerase, which plays a key role in viral RNA transcription. It is feasible that this may reduce antigen formation from retained viral RNA fragments even in the absence of viral replication. This gives it a biologically plausible mechanism for reducing inflammatory burden, where other antivirals, dependent on preventing whole viral replication, have failed.

### Limitations

This study has several important limitations. First, as an open-label, single-arm feasibility trial without a comparator group, the observed changes in clinical, functional, and biomarker measures cannot be solely attributed to remdesivir. Improvements could reflect natural fluctuation in symptom burden, behavioural modification associated with trial participation, or expectation effects. Given the duration and episodic nature of LC, fluctuations in symptom prevalence and severity are unlikely to explain these observations [40]. The Hawthorne effect [41], of trial participation, however, is well established, particularly for conditions that have faced significant stigma in the absence of diagnostic tests [42]. The potential influence of trial participation effects should also be considered in the context of findings from other LC intervention studies. For example, the recent STIMULATE-ICP trial reported improvements in fatigue among participants receiving usual care/no study medication, highlighting the importance of expectation and participation effects in LC research. However, despite similar baseline fatigue severity, the magnitude of improvement in FAS scores observed in the present study appears greater than that reported in the comparator arm of STIMULATE-ICP [33]. While cross-study comparisons should be interpreted cautiously owing to differences in study design, populations, and follow-up duration, these findings suggest that the observed improvements may not be entirely attributable to trial participation effects alone. Nevertheless, the absence of a randomised control group precludes causal inference, and placebo-controlled trials remain essential to establish efficacy.

Second, the relatively short follow-up period precludes assessment of the durability of response. Third, participants were predominantly white and largely recruited from specialist LC clinics, which may limit generalisability to more diverse or community-managed populations. In addition, individuals considered to be at high risk of PEM were excluded because of concerns regarding the burden and potential risks associated with exertion-based assessments. Indeed, 11 of the 20 participants deemed ineligible during screening were excluded on this basis, highlighting the substantial representation of this phenotype within the broader Long COVID population. Consequently, our findings may not be generalisable to individuals with an ME/CFS-like phenotype of LC, a subgroup that experiences substantial functional impairment and may have distinct treatment responses. The effects of remdesivir in individuals with more severe PEM therefore remain uncertain and warrant investigation using study designs that minimise participant burden while maintaining robust outcome assessment. Although the 6MWT and CPET provided important information regarding feasibility and physiological outcomes in the current study, future trials should consider whether these assessments are required for all participants or whether alternative outcome measures, such as patient-reported outcomes, wearable-device data, activity monitoring, and molecular biomarkers, could provide robust efficacy data while enabling broader inclusion of individuals with PEM. A stratified or nested assessment approach may permit detailed physiological evaluation in a subset of participants while reducing barriers to participation and improving generalisability.

Finally, although the study was adequately powered to assess feasibility and the prespecified CPET endpoint, it was not designed to evaluate clinical efficacy, optimise dosing strategies, or identify predictors of treatment response. Future research should therefore prioritise adequately powered randomised controlled trials to determine the efficacy, safety, durability, and cost-effectiveness of remdesivir in LC. Parallel development of diagnostic, physiological, and molecular biomarkers capable of distinguishing likely responders from non-responders will also be important to support more precise treatment approaches.

## Conclusion

We have demonstrated, for the first time, that intravenous administration of remdesivir is feasible for individuals living with LC, with no unexpected safety signals, offering early indications of potential clinical benefit across multiple symptom domains. While these findings are encouraging, they remain preliminary and must be interpreted within the constraints of a single-arm feasibility design. Collectively, these research priorities form the foundation needed to clarify remdesivir’s therapeutic role and determine its potential integration into broader clinical pathways for LC management.

## Data Availability

All data produced in the present study are available upon reasonable request to the authors.

## Declaration of Interests

This study was supported by an unrestricted investigator-sponsored research grant from Gilead Sciences (#IN-UK-540-6640).

## Acknowledgements

The authors would like to acknowledge the tireless work and energy provided by our PPIE group. They have trusted us with this project and, through our ongoing collaboration, have enabled us to deliver this trial. There is no research without you. We would also like to acknowledge colleagues at the University Hospitals Derby and Burton, the Exeter Clinical Research Facility and the University of Derby for their support in the delivery of the trial. Finally, we would like to acknowledge colleagues at the Derby Clinical Trials Support Unit for their support in the initial stages and development of the trial.

## Authorship contributions

MF and JC led the development of the manuscript. MF and TB were co-chief investigators. MF, RA, TB, DS and KK conceptualised the research project, applied for research funding and acted as site leads for the project. RO, CT, SK, and KM led the data collection with support from the lead investigators. Trial management was provided by HN, KAS and the wider Peninsula Clinical Trials Team. KK and HR led the PET-CT imaging. Data management was led by PA and HH, and statistical analysis was led by JC with support from AB and VA. JC and AB had full access to the data in the study and took responsibility for the accuracy of the data analysis. IM led pharmacy aspects, including supporting the development of the pharmacy protocol across sites. LS was the lead patient and public involvement and engagement representative and was supported by a wider PPIE network. All authors read and approved the final manuscript before submission.

## Study Funding

Provided by an unrestricted investigator-sponsored research grant from Gilead Sciences (#IN-UK-540-6640).

## APC Funding

APC Funding is provided via Loughborough University’s Open Access Agreements.

## Conflicts of Interest

None

## Data availability

The datasets used and/or analysed during the current study are available from the corresponding author on reasonable request.

## Supplementary table

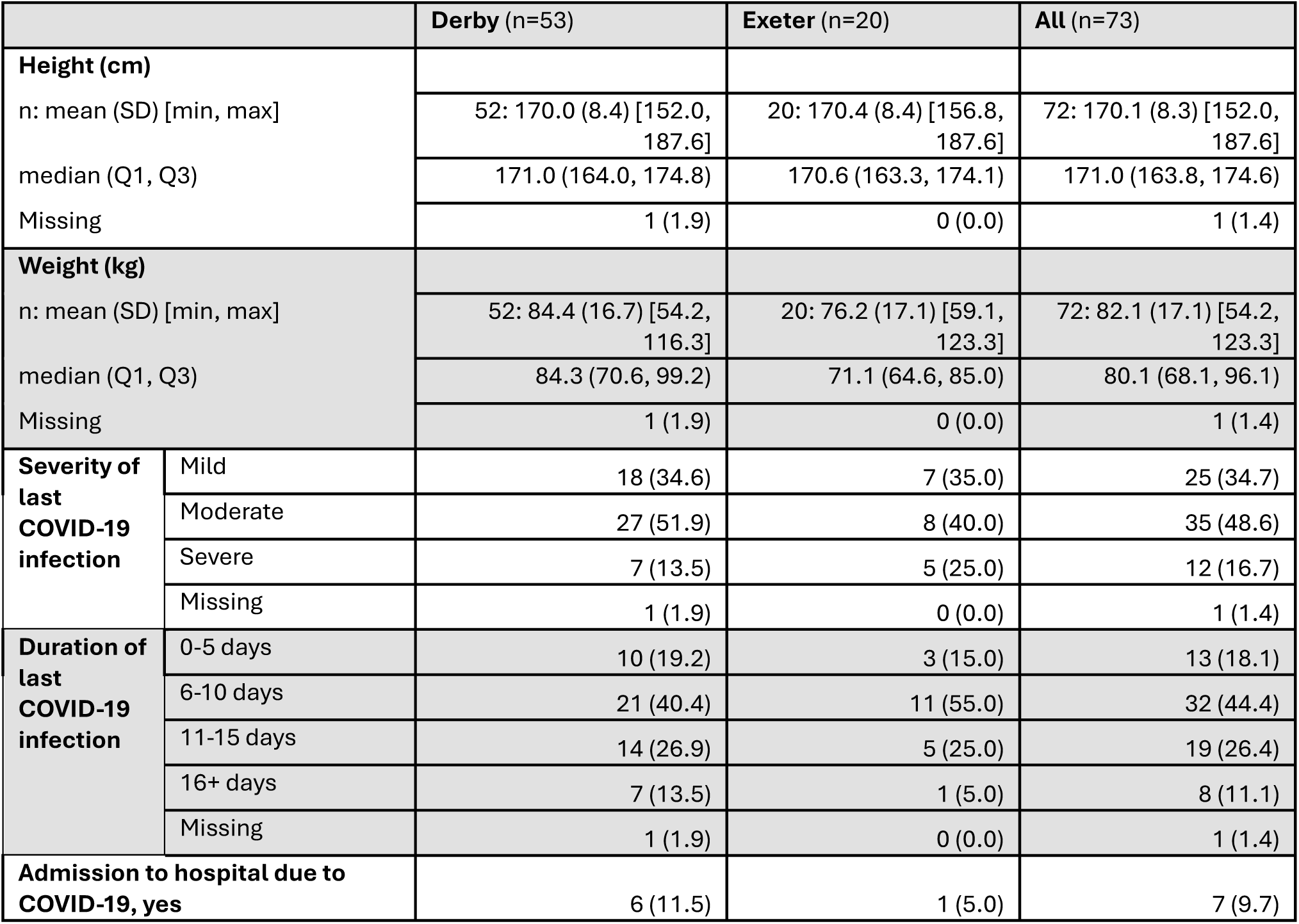
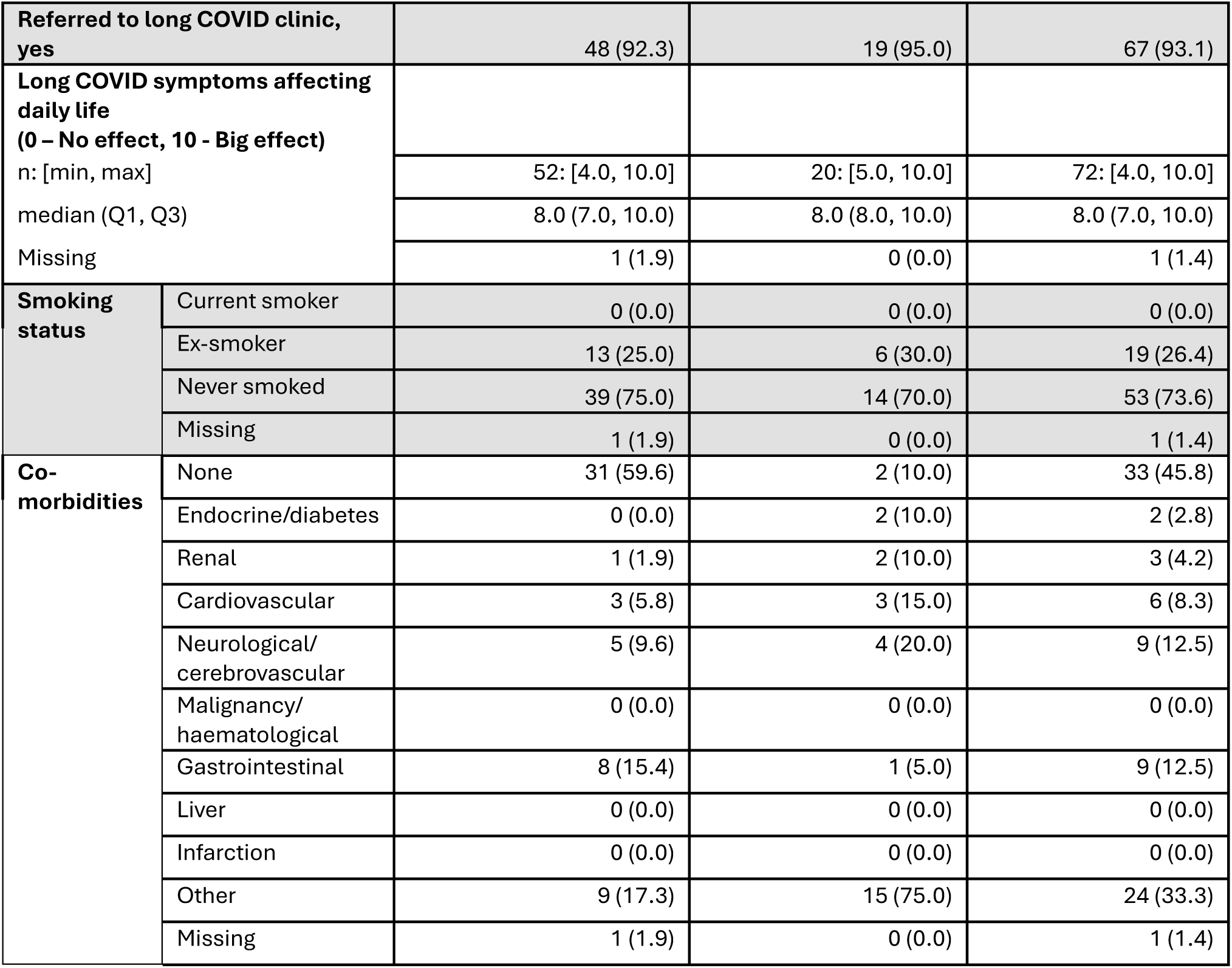

## Notes

### Competing Interest Statement

The authors have declared no competing interest.

### Clinical Trial

NCT05911906

### Clinical Protocols

https://link.springer.com/article/10.1186/s40814-026-01823-9

### Author Declarations

Ethics approval was obtained from the UK Health Research Authority Research Ethics Committee (24/SC/0118), the Administration of Radioactive Substances Advisory Committee (AA-8765), and the Medicines and Healthcare products Regulatory Agency (24/SC/0118).

